# Changes in Cardiorespiratory Fitness in Patients with Human Papillomavirus (HPV)-Related Oropharyngeal Cancer Undergoing Chemoradiotherapy

**DOI:** 10.64898/2026.04.03.26350101

**Authors:** Michael Burgess, Jack Thomson, Benjamin Fox, Emili Salas Diaz, Guy S Taylor, Callum G Brownstein, Muhammad Shahid Iqbal, James O’Hara, Rhona C F Sinclair, Samuel T Orange

## Abstract

**Purpose:** Chemoradiotherapy (CRT) for human papillomavirus-related oropharyngeal cancer (HPV+ OPC) causes substantial treatment-related toxicity, with well-known adverse effects on quality of life (QoL), weight loss, and self-reported physical functioning. However, its impact on objectively measured cardiorespiratory fitness is unknown. This study examined changes in cardiorespiratory fitness, body composition, grip strength, and patient-reported outcomes in patients with HPV+ OPC undergoing CRT.

**Methods:** We invited 20 patients with HPV+ OPC scheduled for CRT (age: 61.2±7.1 years, female: n=4) to complete assessments at three timepoints: pre-CRT (baseline), 2-weeks post-CRT, and 8-weeks post-CRT. Cardiorespiratory fitness was assessed using a maximal incremental cardiopulmonary exercise test (CPET). Body composition was estimated using segmental bioelectrical impedance analysis. QoL was assessed using the EORTC QLQ-C30 and QLQ-H&N43, and physical activity was self-reported using the International Physical Activity Questionnaire-Short Form. The primary outcome was change in oxygen consumption at the anaerobic threshold (V̇O₂ at AT) measured during CPET; an objective, effort-independent marker of cardiorespiratory fitness.

**Results:** Mean V̇O₂ at AT declined from 16.0±3.8 ml/kg/min at baseline to 12.0±3.4 ml/kg/min at 2-weeks post-CRT (adjusted mean change: −4.2, 95% CI: −5.4 to −3.0 ml/kg/min) and remained low at 8-weeks post-CRT. Peak oxygen consumption (V̇O_2_peak: −7.4, −9.3 to −5.4 ml/kg/min), body mass (−8.5, −10.7 to −6.2 kg), fat-free mass (−6.4, −7.7 to −5.0 kg), grip strength (−4.1, −7.2 to −0.99 kg), global health status (−26.9, −39.2 to −14.6 points), fatigue (49.8, 33.7 to 65.8 points), and several disease-specific symptoms were also adversely affected at 2-weeks post-CRT and remained impaired at 8 weeks.

**Conclusion:** This is the first study to estimate the impact of CRT on cardiopulmonary fitness in patients with HPV+ OPC. Cardiorespiratory fitness declined by ∼25% following CRT and remained reduced at 8-weeks. Targeted interventions to mitigate these adverse physiological effects warrants further investigation.

## INTRODUCTION

The incidence of oropharyngeal cancer related to human papilloma virus (HPV+ OPC) is increasing in many Western countries (1). Standard treatment for locally advanced HPV+ OPC involves six weeks of combined radiotherapy and chemotherapy (chemoradiotherapy; CRT) (2). Although three-year survival rates exceed 80%, CRT is associated with substantial local and systemic toxicity (3).

Patients commonly experience clinically important weight loss, fatigue, and declines in QoL following treatment, despite multidisciplinary care and personalised nutritional support (4,5). No study has objectively characterised changes in cardiorespiratory fitness following CRT in patients with HPV+ OPC, despite it being strongly and independently associated with morbidity and mortality in people with cancer (6).

Cardiorespiratory fitness can be objectively assessed using cardiopulmonary exercise testing (CPET), which involves the collection of respired gases during a maximal incremental exercise test. CPET provides an integrated assessment of cardiovascular, pulmonary, and musculoskeletal responses to increasing physiological demand. Key CPET-derived variables include peak oxygen consumption (V̇O₂ peak), an index of peak cardiorespiratory fitness, and oxygen consumption at the anaerobic threshold (V̇O_2_ at AT), which reflects the transition to greater reliance on anaerobic metabolism. V̇O_2_ at AT is an objective, submaximal, effort-independent marker of change across integrated physiological systems (7). Assessment of these parameters before and after CRT may improve understanding of the physiological impact of treatment and inform the design of targeted interventions.

The aim of this study was to examine changes in cardiorespiratory fitness alongside body composition, grip strength, and patient-reported outcomes in patients with HPV+ OPC undergoing CRT.

## METHODS

### Study design

This was a prospective, single-center, observational study conducted between April 2024 and September 2025 in Newcastle upon Tyne, UK. Ethical approval was obtained from an NHS Research Ethics Committee (24/PR/0040) and the study was prospectively registered (XXX). All participants provided written informed consent.

Eligible patients were identified during multidisciplinary team discussions and approached at outpatient clinics. Inclusion criteria were: (i) aged ≥18 years; (ii) histological diagnosis of HPV+ OPC; and (iii) scheduled to receive CRT. Exclusion criteria were: (i) absolute or relative contraindication to CPET (8); (ii) ECG indicating high-grade arrhythmia or coronary heart disease; (iii) musculoskeletal or neurological conditions limiting exercise.

Participants completed assessments at three time-points: pre-CRT (Visit 1; baseline); 2-weeks post-CRT (Visit 2); and 8-weeks post-CRT (Visit 3). Standard CRT comprised 66Gy radiotherapy delivered in 30 fractions (Monday-Friday for 6-weeks) with concomitant cisplatin (weekly 40 mg/m² or 3-weekly 100 mg/m²).

The primary outcome was V̇O_2_ at AT measured during a CPET. Secondary outcomes other CPET-derived indices, body composition, grip strength, hemoglobin, QoL, and self-reported physical activity.

### Study assessments

Participants performed an incremental exercise test to volitional exhaustion on an electronically braked cycle ergometer (VIAsprint 200P, Ergoline GmbH) following a standardised ramp protocol (7). Breath-by-breath data (Cortex MetaLyzer 3B), heart rate (Polar H10), and perceived exertion (6-20 Borg scale) were recorded throughout.

Breath-by-breath data were averaged using a moving average (middle 5 of 7 breaths). V̇O_2_ at AT was independently estimated by two analysts based on the inflection point in carbon dioxide output (V CO_2_), ventilatory equivalents, and end-tidal pressures for O_2_ and CO_2_, with ≥7.5% discrepancies resolved through discussion. Additional CPET variables included V̇O_2_peak, peak O_2_ pulse, respiratory exchange ratio (RER), peak ventilation (V̇E), V̇E/ V̇CO_2_ slope, O_2_ uptake efficiency slope (OUES), and the V̇O_2_-work rate relationship (ΔV̇O_2_/ΔWR).

Body mass, fat mass, fat-free mass, and skeletal muscle mass were estimated by validated, multi-frequency, segmental bioelectrical impedance analysis (Seca mBCA 515) (9). Maximal isometric grip strength (dominant hand) was measured using a hand-held digital grip dynamometer (Takei Scientific Instruments). Spirometry assessed forced expiratory volume in 1 s (FEV□), forced vital capacity (FVC), and peak expiratory flow.

Cancer-related and disease-specific QoL was assessed using the EORTC QLQ-C30 and EORTC QLQ–H&N43 questionnaires, respectively (10,11). The International Physical Activity–Short Form (IPAQ-SF) assessed self-reported physical activity (12).

### Statistical analysis

We aimed to recruit 30 participants during a planned 14-month recruitment period, accounting for 20% dropout, as this sample size would provide 80% power (α = 0.05) to detect a change in V̇O_2_ at AT of ≥1.5±2.5 ml/kg/min (13,14). Twenty participants were recruited within the recruitment window.

Multilevel mixed-effects models assessed changes over time, with timepoint as a fixed effect and participant ID as a random effect to account for repeated measures. Models were fit using maximum likelihood estimation. Adjusted mean changes (estimated marginal means) with 95% confidence intervals and Holm-corrected p-values are presented. Statistical significance was set at p<0.05. Analyses were performed in R version 4.4.3.

## RESULTS

### Participant characteristics

Twenty patients with HPV+ OPC (T1-4, N1-3, M0) were recruited (**Table 1**). Six and three participants did not attend follow-up assessments at 2- and 8-weeks post-CRT, respectively, due to hospitalisation, treatment-related side effects, or being uncontactable (**FigA1**). Actual timing of post-CRT assessments was 2.9±0.9 weeks and 8.8±2.0 weeks, respectively.

**Table 1.** Participant characteristics at baseline (n=20)

| | Mean ± SD<br>or number<br>(%) |
| --- | --- |
| Age (years) | 61.2 ± 7.1 |
| Female | 4 (20%) |
| BMI (kg/m<sup>2</sup>) | 30.2 ± 3.8 |
| Body mass (kg) | 88.0 ± 13.2 |
| Fat mass (%) | 35.1 ± 6.8 |
| V̇O<sub>2peak</sub> (ml/kg/min) | 26.5 ± 6.0 |
| Systolic blood pressure (mmHg) | 133 ± 13.8 |
| Diastolic blood pressure (mmHg) | 83.1 ± 8.2 |
| Self-reported alcohol intake (units/week) | 10.0 ± 11.9 |
| **Ethnicity** |  |
| White British | 19 (95%) |
| Black African | 1 (5%) |
| **Smoking status** |  |
| Current smoker | 0 (0%) |
| Previous smoker | 9 (45%) |
| **Comorbidities** |  |
| Cardiovascular disease | 4 (20%) |
| Type II diabetes | 3 (15%) |
| Musculoskeletal disorder | 4 (20%) |
| Other | 4 (20%) |
| None | 7 (35%) |
| **Chemotherapy schedule** |  |
| Weekly | 17 (85%) |
| 3-Weekly | 3 (15%) |

BMI: body mass index, V̇O<sub>2peak</sub>: peak oxygen
uptake.

**Fig 1.**
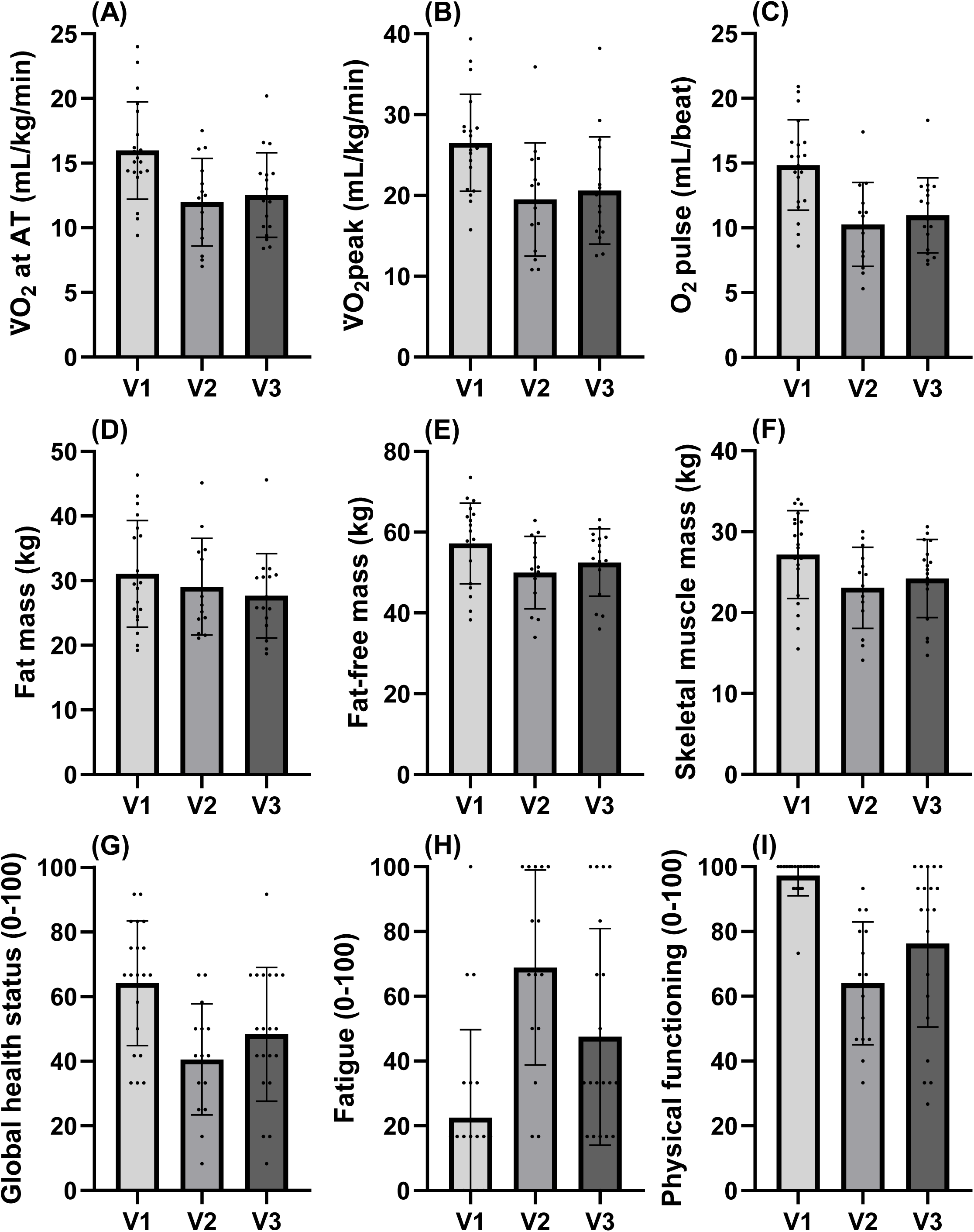
Changes in cardiorespiratory fitness (panels A-C), body composition (D-F), and quality of life (G-I) from baseline (Visit 1) to 2-weeks (Visit 2) and 8-weeks (Visit 3) post-chemoradiotherapy. O_2_ pulse: oxygen pulse, V̇O_2_ at AT: oxygen uptake at the ventilatory anaerobic threshold, V̇O_2_peak: peak oxygen uptake.

### Cardiorespiratory fitness

Mean V̇O₂ at AT declined from 16.0±3.8 ml/kg/min at baseline to 12.0±3.4 ml/kg/min at 2-weeks post-CRT (adjusted mean change: −4.2, 95% CI: −5.4 to −3.0 ml/kg/min) and remained reduced at 8-weeks (**Table 2**). V̇O_2_peak (−7.4, −9.3 to −5.4 mL/kg/min), peak O_2_ pulse (−4.1, −4.9 to −3.3 mL/beat), and OUES (−0.79, −0.93 to −0.64) were also reduced at 2-weeks and remained low at 8-weeks post-CRT. Peak heart rate and respiratory exchange ratio did not significantly differ across visits and no adverse events during CPET were reported. Descriptive statistics for all outcomes are displayed in **Table A1** and adjusted mean changes in **Table A2**.

**Table 2.**
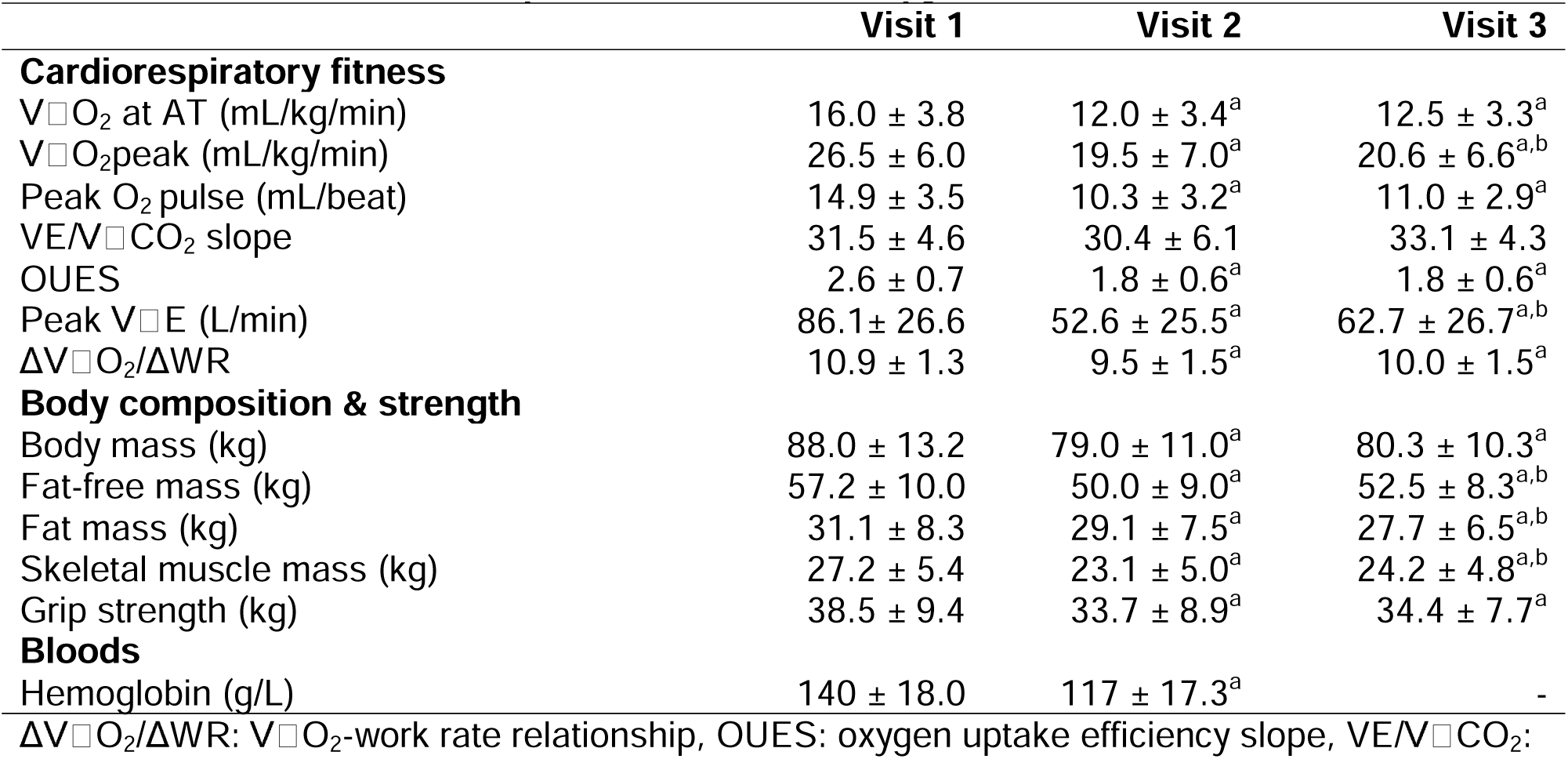
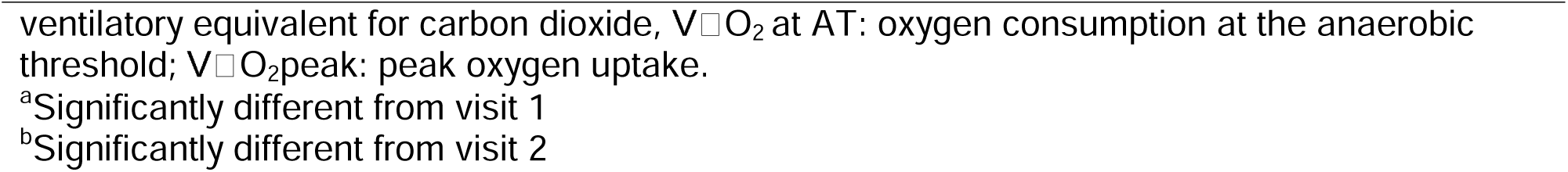
Cardiorespiratory fitness and body composition at baseline (Visit 1), 2-weeks (Visit 2) and 8-weeks (Visit 3) post-chemoradiotherapy (mean ± SD)

|  | Visit 1 | Visit 2 | Visit 3 |
| --- | --- | --- | --- |
| **Cardiorespiratory fitness** |  |  |  |
| V$\dot{V}$O<sub>2</sub> at AT (mL/kg/min) | 16.0 ± 3.8 | 12.0 ± 3.4<sup>a</sup> | 12.5 ± 3.3<sup>a</sup> |
| V$\dot{V}$O<sub>2</sub>peak (mL/kg/min) | 26.5 ± 6.0 | 19.5 ± 7.0<sup>a</sup> | 20.6 ± 6.6<sup>a,b</sup> |
| Peak O<sub>2</sub> pulse (mL/beat) | 14.9 ± 3.5 | 10.3 ± 3.2<sup>a</sup> | 11.0 ± 2.9<sup>a</sup> |
| VE/V$\dot{V}$CO<sub>2</sub> slope | 31.5 ± 4.6 | 30.4 ± 6.1 | 33.1 ± 4.3 |
| OUES | 2.6 ± 0.7 | 1.8 ± 0.6<sup>a</sup> | 1.8 ± 0.6<sup>a</sup> |
| Peak V$\dot{V}$E (L/min) | 86.1 ± 26.6 | 52.6 ± 25.5<sup>a</sup> | 62.7 ± 26.7<sup>a,b</sup> |
| ΔV$\dot{V}$O<sub>2</sub>/ΔWR | 10.9 ± 1.3 | 9.5 ± 1.5<sup>a</sup> | 10.0 ± 1.5<sup>a</sup> |
| **Body composition & strength** |  |  |  |
| Body mass (kg) | 88.0 ± 13.2 | 79.0 ± 11.0<sup>a</sup> | 80.3 ± 10.3<sup>a</sup> |
| Fat-free mass (kg) | 57.2 ± 10.0 | 50.0 ± 9.0<sup>a</sup> | 52.5 ± 8.3<sup>a,b</sup> |
| Fat mass (kg) | 31.1 ± 8.3 | 29.1 ± 7.5<sup>a</sup> | 27.7 ± 6.5<sup>a,b</sup> |
| Skeletal muscle mass (kg) | 27.2 ± 5.4 | 23.1 ± 5.0<sup>a</sup> | 24.2 ± 4.8<sup>a,b</sup> |
| Grip strength (kg) | 38.5 ± 9.4 | 33.7 ± 8.9<sup>a</sup> | 34.4 ± 7.7<sup>a</sup> |
| **Bloods** |  |  |  |
| Hemoglobin (g/L) | 140 ± 18.0 | 117 ± 17.3<sup>a</sup> | - |

ΔV $\dot{V}$ O<sub>2</sub>/ ΔWR: V $\dot{V}$ O<sub>2</sub>-work rate relationship, OUES: oxygen uptake efficiency slope, VE/V $\dot{V}$ CO<sub>2</sub>:
ventilatory equivalent for carbon dioxide, $V\dot{V}O_2$ at AT: oxygen consumption at the anaerobic threshold; $V\dot{V}O_{2peak}$ : peak oxygen uptake.
<sup>a</sup>Significantly different from visit 1
<sup>b</sup>Significantly different from visit 2

**Table 3.** Self-reported outcomes at baseline (Visit 1) to 2-weeks (Visit 2) and 8-weeks (Visit 3) post-chemoradiotherapy (mean ± SD)

|  | Visit 1 | Visit 2 | Visit 3 |
| --- | --- | --- | --- |
| **Physical activity** |  |  |  |
| Walking (min/week) | 513 ± 354 | 187 ± 247<sup>a</sup> | 410 ± 290<sup>b</sup> |
| MVPA (min/week) | 236 ± 419 | 71.0 ± 147 | 168 ± 326 |
| Total PA (min/week) | 748 ± 533 | 258 ± 339<sup>a</sup> | 578 ± 434<sup>b</sup> |
| **QLQ-C30** |  |  |  |
| Global Health Status | 64.2 ± 19.3 | 40.6 ± 17.2<sup>a</sup> | 48.3 ± 20.7<sup>a,b</sup> |
| Fatigue | 22.5 ± 27.2 | 68.9 ± 30.1<sup>a</sup> | 47.5 ± 33.5<sup>a,b</sup> |
| Physical Functioning | 97.3 ± 6.3 | 64.0 ± 19.0<sup>a</sup> | 76.3 ± 25.8<sup>a</sup> |
| Role Functioning | 89.2 ± 21.8 | 28.9 ± 31.2<sup>a</sup> | 54.2 ± 35.4<sup>a,b</sup> |
| Emotional Functioning | 78.8 ± 21.0 | 70.0 ± 14.7 | 71.2 ± 27.8 |
| Cognitive Functioning | 86.7 ± 21.4 | 68.9 ± 27.4<sup>a</sup> | 69.2 ± 29.3 |
| Social Functioning | 81.7 ± 24.7 | 43.3 ± 27.3<sup>a</sup> | 53.3 ± 34.9<sup>a,b</sup> |
| **H&N43** |  |  |  |
| Problems opening mouth | 6.7 ± 17.4 | 44.4 ± 34.9<sup>a</sup> | 26.7 ± 27.8<sup>a,b</sup> |
| Problem with senses | 6.7 ± 19.8 | 47.8 ± 19.8<sup>a</sup> | 35.0 ± 31.0<sup>a,b</sup> |
| Swallowing | 9.6 ± 22.2 | 56.7 ± 29.9<sup>a</sup> | 36.3 ± 34.0<sup>a,b</sup> |
| Speech | 10.3 ± 19.9 | 47.6 ± 38.3<sup>a</sup> | 40.0 ± 30.8<sup>a,b</sup> |
| Weight loss | 6.7 ± 13.7 | 31.1 ± 19.8<sup>a</sup> | 28.3 ± 32.9<sup>a</sup> |

H&N43: Head and Neck Module 43 items, MVPA: moderate- to vigorous-intensity physical activity (e.g., bicycling, tennis, aerobics), QLQ-C30: Quality of Life Questionnaire Core 30.
<sup>a</sup>Significantly different from visit 1
<sup>b</sup>Significantly different from visit 2

### Body composition

Reductions in total body mass (−8.5, −10.7 to −6.2 kg), fat mass (−2.2, −3.7 to −0.69 kg), fat-free mass (−6.4, −7.7 to −5.0 kg), and skeletal muscle mass (−3.6, −4.5 to −2.7 kg) were observed 2-weeks post-CRT and persisted 8-weeks post-CRT. Critical weight loss (>5% total body mass loss) was observed in 86% of participants. Fat-free mass and skeletal muscle mass showed a small increase from 2-weeks to 8-weeks post-CRT (**Table 2**).

### Self-reported outcomes

Global health status (−26.9, −39.2 to −14.6 points), fatigue (+49.8, 33.7 to 65.8 points), and several QoL domains and cancer-specific symptoms were adversely affected at 2-weeks post-CRT (**Table 2**). Most self-reported outcomes remained impaired at 8-weeks, although global health status, fatigue, and specific symptoms improved between 2-weeks to 8-weeks post-CRT. Time spent doing physical activities declined at 2-weeks and increased towards baseline by 8-weeks post-CRT.

## DISCUSSION

This is the first study to objectively measure changes in cardiorespiratory fitness in patients with HPV+ OPC undergoing CRT. Our main finding was that V̇O₂ at AT declined by ∼25% following CRT and remained impaired at 8-weeks post-treatment. These findings demonstrate the substantial physiological impact of CRT and highlight the need for targeted interventions.

The reduction in oxygen uptake (V̇O_2_) at AT reflects an earlier transition to reliance on anaerobic metabolism during incremental exercise. Lower V̇O_2_ at AT, together with the decrease in V̇O_2_peak, indicate a marked impairment in oxygen delivery and/or utilisation. The magnitude of reduction in cardiorespiratory fitness observed is substantially larger than that reported in other cancer populations; the adjusted mean change in V̇O₂ at AT in our cohort was −4.2 ml/kg/min, whereas previous studies in patients with esophagogastric cancer undergoing chemotherapy have reported mean reductions of ∼1.5-2.5 ml/kg/min (13,14). The larger decline in our study may partly reflect the additional toxic effects of radiotherapy alongside cisplatin chemotherapy (15).

Low V̇O₂ peak post-cancer diagnosis is associated with higher symptom burden (16) and greater risk of all-cause, cardiovascular, and cancer mortality (6). Patients in our study had normal cardiorespiratory fitness prior to treatment; mean baseline V̇O₂ peak (26.5 ml/kg/min) corresponds to 102% of the predicted value based on age and sex-matched reference values for the general population (17). Despite normal baseline fitness, the substantial treatment-related reduction in cardiorespiratory fitness (if sustained long-term) may have implications for future health outcomes.

Our findings highlight the need for evidence-based strategies aimed at mitigating the adverse physiological effects of CRT, such as targeted physical activity interventions, which hold promise for improving cardiorespiratory fitness and longer-term outcomes in people receiving systemic cancer therapy (18). Further work is required to understand how physical activity interventions may be implemented within standard care pathways for patients with HPV+ OPC, who face distinct barriers to physical activity, including having a dry throat and an intensive treatment regimen (19). Additionally, our data may provide a benchmark for interpreting outcomes in future real-world observational or intervention trials.

Although self-reported time spent doing physical activities declined during CRT, physical activity levels (principally walking) returned close to baseline by 8-weeks post-CRT. Given cardiorespiratory fitness remained suppressed at 8-weeks, these findings tentatively suggest walking alone may be insufficient to restore oxidative capacity following CRT and more targeted exercise interventions may be required. Alternatively, it may require a sustained period of activity post-CRT to restore cardiorespiratory fitness.

The mechanisms underlying reduced cardiorespiratory fitness following CRT are likely multifactorial. Treatment-related cellular toxicity may disrupt several physiological systems that contribute to oxygen transport and utilisation, as well as having indirect effects on physical activity and nutritional intake (20). Adverse changes across multiple CPET indices, together with reductions in haemoglobin concentration and skeletal muscle mass, indicate impairments in both oxygen delivery and peripheral oxygen utilisation. The absence of change in the V̇E/V̇CO₂ slope and preserved breathing reserve suggest that ventilatory limitation may not be an underlying factor. Future studies using additional physiological assessments, such as exercise echocardiography and near-infrared spectroscopy, may help further delineate the underlying mechanisms.

Our findings demonstrate adverse changes in body weight, global health status, and fatigue following CRT, consistent with previous studies (4,5). Approximately 75% of weight lost at 2-weeks post-CRT comprised fat-free mass, which may increase long-term risk of muscle-related health conditions.

A limitation of this study is missing follow-up data due to participant hospitalisation or treatment-related illness, which may have led to underestimation of the true effects. Additionally, the observational design precludes causal inference, although the magnitude of consistency of the changes during CRT are compelling.

In conclusion, cardiorespiratory fitness declined by ∼25% following CRT in patients with HPV+ OPC and remained impaired at 8-weeks post treatment. Targeted interventions to mitigate these adverse physiological effects warrants further investigation.

## Supporting information

FigA1

## Data Availability

Anonymized individual-level research data and statistical code are available on the Open Science Framework (https://osf.io/rk83d/)

