## Supplementary material for "Changes in Cardiorespiratory Fitness in Patients with Human Papillomavirus (HPV)-Related Oropharyngeal Cancer Undergoing Chemoradiotherapy": Table A1

**Table A2. Adjusted mean changes in cardiorespiratory fitness, body composition, hemoglobin, and self-reported outcomes at baseline (Visit 1), 2-weeks (Visit 2) and 8-weeks (Visit 3) post-chemoradiotherapy (adjusted mean change, 95% CI, Holm-corrected p-value)**

| | Visit 2 – Visit 1 | Visit 3 - Visit 1 | Visit 3 - Visit 2 |
|---|---|---|---|
| **Cardiorespiratory fitness** | | | |
| $\dot{V}O_2$ at AT (L/min) | -473 (-587, -359), p = <0.001 | -403 (-545, -261), p = <0.001 | 70 (-17.1, 157), p = 0.111 |
| $\dot{V}O_2$ at AT (mL/kg/min) | -4.2 (-5.37, -3.02), p = <0.001 | -3.43 (-4.79, -2.08), p = <0.001 | 0.763 (-0.401, 1.93), p = 0.190 |
| $\dot{V}O_2$peak (L/min) | -857 (-1080, -639), p = <0.001 | -664 (-872, -456), p = <0.001 | 193 (82.6, 304), p = 0.001 |
| $\dot{V}O_2$peak (mL/kg/min) | -7.39 (-9.34, -5.44), p = <0.001 | -5.51 (-7.44, -3.58), p = <0.001 | 1.88 (0.317, 3.45), p = 0.020 |
| Peak $O_2$ pulse (mL/beat) | -4.13 (-4.91, -3.34), p = <0.001 | -3.8 (-4.85, -2.75), p = <0.001 | 0.326 (-0.814, 1.47), p = 0.563 |
| Peak RER | -0.016 (-0.056, 0.026), p = 1.000 | -0.01 (-0.046, 0.022), p = 1.000 | 0.00 (-0.03, 0.040), p = 1.000 |
| Peak $\dot{V}E$ (L/min) | -37.2 (-49.4, -25), p = <0.001 | -23.8 (-34.4, -13.1), p = <0.001 | 13.4 (7.92, 19), p = <0.001 |
| Breathing reserve (%) | 28.4 (15.6, 41.2), p = <0.001 | 25.9 (15.6, 36.1), p = <0.001 | -2.55 (-13.3, 8.18), p = 0.630 |
| VE/$\dot{V}CO_2$ slope | -1.09 (-4.77, 2.59), p = 0.549 | 1.55 (-0.268, 3.38), p = 0.275 | 2.64 (-0.716, 6), p = 0.275 |
| $\Delta\dot{V}O_2/\Delta WR$ | -1.3 (-1.91, -0.694), p = <0.001 | -0.881 (-1.63, -0.138), p = 0.044 | 0.423 (-0.354, 1.2), p = 0.275 |
| OUES | -0.79 (-0.93, -0.64), p = <0.001 | -0.81 (-1.0, -0.59), p = <0.001 | -0.0248 (-0.22, 0.17), p = 0.794 |
| Chronotropic index | -0.067 (-0.12, -0.011), p = 0.064 | -0.05 (-0.11, 0.010), p = 0.203 | 0.0174 (-0.05, 0.080), p = 0.573 |
| Peak HR (bpm) | -10.8 (-19.7, -1.84), p = 0.059 | -8.13 (-17.2, 0.909), p = 0.152 | 2.67 (-6.98, 12.3), p = 0.576 |
| Peak RPE | -0.0112 (-1.19, 1.17), p = 0.985 | 0.952 (-0.177, 2.08), p = 0.191 | 0.963 (0.249, 1.68), p = 0.030 |
| FEV1 (L) | -0.14 (-0.36, 0.076), p = 0.392 | 0.0511 (-0.07, 0.17), p = 0.401 | 0.191 (-0.06, 0.44), p = 0.385 |

| | | | |
|---|---|---|---|
| FVC (L) | -0.231 (-0.545, 0.0829), p = 0.302 | 0.0355 (-0.138, 0.209), p = 0.678 | 0.267 (-0.0549, 0.588), p = 0.302 |
| FEV1/FVC | 0.003 (-0.02, 0.02), p = 1.000 | 0.00 (-0.03, 0.03), p = 1.000 | -0.00 (-0.0183, 0.0133), p = 1.000 |
| PEF (L/min) | -76.7 (-111, -42.2), p = <0.001 | -15.9 (-61.6, 29.7), p = 0.482 | 60.7 (23.2, 98.3), p = 0.005 |
| **Body composition** | | | |
| BMI (kg/m2) | -2.91 (-3.7, -2.12), p = <0.001 | -2.87 (-3.91, -1.82), p = <0.001 | 0.0442 (-0.559, 0.648), p = 0.882 |
| Body mass (kg) | -8.47 (-10.7, -6.24), p = <0.001 | -8.46 (-11.5, -5.39), p = <0.001 | 0.00841 (-1.78, 1.8), p = 0.992 |
| Fat-free mass (kg) | -6.37 (-7.71, -5.03), p = <0.001 | -4.85 (-6.17, -3.53), p = <0.001 | 1.52 (0.622, 2.41), p = 0.002 |
| Fat-free mass (%) | -1.19 (-2.22, -0.16), p = 0.050 | 1.07 (-0.715, 2.85), p = 0.230 | 2.26 (0.905, 3.61), p = 0.006 |
| Fat mass (kg) | -2.19 (-3.69, -0.689), p = 0.012 | -4.01 (-6.55, -1.46), p = 0.010 | -1.82 (-3.54, -0.0942), p = 0.039 |
| Fat mass (%) | 1.13 (0.0569, 2.2), p = 0.079 | -1.07 (-2.85, 0.715), p = 0.230 | -2.19 (-3.64, -0.752), p = 0.013 |
| Skeletal muscle mass (kg) | -3.63 (-4.51, -2.74), p = <0.001 | -3 (-3.88, -2.12), p = <0.001 | 0.63 (0.153, 1.11), p = 0.011 |
| Waist circumference (cm) | -2.67 (-6.65, 1.32), p = 0.364 | -3.88 (-7.82, 0.0568), p = 0.160 | -1.21 (-3.61, 1.18), p = 0.364 |
| Hip circumference (cm) | -2.95 (-5.67, -0.225), p = 0.070 | -3.14 (-5.17, -1.11), p = 0.011 | -0.193 (-2.63, 2.24), p = 0.872 |
| Hand grip (kg) | -4.1 (-7.21, -0.987), p = 0.023 | -4.04 (-6.31, -1.77), p = 0.003 | 0.0589 (-1.81, 1.92), p = 0.949 |
| **Bloods** | | | |
| Hemoglobin (g/L) | -22.8 (-30.1, -15.5), p = <0.001 | | |
| **Physical activity** | | | |
| Walking (min/week) | -353 (-526, -180), p = <0.001 | -103 (-283, 77.3), p = 0.253 | 250 (126, 375), p = <0.001 |

| | | | |
|---|---|---|---|
| Moderate (min/week) | -143 (-324, 38.4), p = 0.237 | -22.5 (-139, 94.4), p = 0.698 | 120 (-12.1, 252), p = 0.220 |
| Vigorous (min/week) | -59.3 (-136, 17.5), p = 0.378 | -45 (-137, 46.8), p = 0.651 | 14.3 (-17, 45.6), p = 0.651 |
| MVPA (min/week) | -215 (-467, 37.1), p = 0.184 | -67.5 (-265, 130), p = 0.492 | 147 (8.67, 286), p = 0.114 |
| Total (min/week) | -570 (-897, -243), p = 0.002 | -171 (-458, 117), p = 0.236 | 399 (201, 598), p = <0.001 |
| **QLQ-C30** | | | |
| Global Health Status | -26.9 (-39.2, -14.6), p = <0.001 | -15.8 (-27.9, -3.76), p = 0.023 | 11.1 (2, 20.2), p = 0.023 |
| Physical Functioning | -34.7 (-45, -24.4), p = <0.001 | -21 (-33.4, -8.59), p = 0.003 | 13.7 (-0.41, 27.8), p = 0.057 |
| Role Functioning | -63.4 (-81, -45.7), p = <0.001 | -35 (-54.1, -15.9), p = 0.001 | 28.4 (9.65, 47.1), p = 0.004 |
| Emotional Functioning | -9.79 (-20.7, 1.08), p = 0.228 | -7.5 (-24.2, 9.17), p = 0.734 | 2.29 (-9.29, 13.9), p = 0.734 |
| Cognitive Functioning | -18.2 (-32.7, -3.77), p = 0.045 | -17.5 (-33.9, -1.06), p = 0.075 | 0.731 (-17.5, 19), p = 0.935 |
| Social Functioning | -43.4 (-60.4, -26.3), p = <0.001 | -28.3 (-47.3, -9.38), p = 0.009 | 15 (1.46, 28.6), p = 0.031 |
| Fatigue | 49.8 (33.7, 65.8), p = <0.001 | 25 (3.57, 46.4), p = 0.026 | -24.8 (-44, -5.55), p = 0.026 |
| Nausea and vomiting | 19.9 (5.72, 34.1), p = 0.022 | 15 (1.07, 28.9), p = 0.071 | -4.93 (-22.6, 12.7), p = 0.573 |
| Pain | 23.9 (5.54, 42.2), p = 0.037 | 20 (2.46, 37.5), p = 0.053 | -3.86 (-24.8, 17), p = 0.710 |
| Dyspnoea | 22.8 (13.6, 32.1), p = <0.001 | 21.7 (6.65, 36.7), p = 0.012 | -1.17 (-18.2, 15.8), p = 0.890 |
| Insomnia | -3.8 (-21.1, 13.5), p = 1.000 | 3.33 (-12.2, 18.9), p = 1.000 | 7.13 (-7.25, 21.5), p = 0.962 |
| Appetite loss | 51.1 (36.2, 66.1), p = <0.001 | 28.3 (10.4, 46.3), p = 0.006 | -22.8 (-41.5, -4.09), p = 0.018 |
| Constipation | 24 (7.12, 40.8), p = 0.020 | 20 (3.35, 36.7), p = 0.040 | -3.98 (-18.6, 10.6), p = 0.582 |
| Diarrhoea | 18.3 (8.86, 27.8), p = 0.001 | 18.3 (4.85, 31.8), p = 0.018 | 0.0165 (-13.7, 13.8), p = 0.998 |

| | | | |
|---|---|---|---|
| Financial difficulty | 4.08 (-7.31, 15.5), p = 0.673 | 10 (-6.43, 26.4), p = 0.673 | 5.92 (-5.02, 16.9), p = 0.673 |
| **H&N43** | | | |
| Pain in the mouth | 35.1 (18.7, 51.6), p = <0.001 | 20 (2.55, 37.5), p = 0.026 | -15.1 (-23, -7.3), p = <0.001 |
| Swallowing | 55.5 (36.6, 74.5), p = <0.001 | 26.7 (8.07, 45.3), p = 0.006 | -28.9 (-43.1, -14.7), p = <0.001 |
| Problems with teeth | 3.63 (-9.17, 16.4), p = 0.568 | 13.3 (-0.381, 27), p = 0.169 | 9.7 (-0.795, 20.2), p = 0.169 |
| Dry mouth and sticky saliva | 64.8 (53.9, 75.8), p = <0.001 | 52.5 (39.2, 65.8), p = <0.001 | -12.3 (-19.8, -4.88), p = 0.002 |
| Problems with senses | 40.7 (29.8, 51.6), p = <0.001 | 28.3 (17, 39.7), p = <0.001 | -12.4 (-23.4, -1.3), p = 0.030 |
| Speech | 48.9 (26.6, 71.2), p = <0.001 | 29.7 (15.2, 44.1), p = <0.001 | -19.3 (-35.4, -3.12), p = 0.021 |
| Body Image | 28.1 (15.2, 41), p = <0.001 | 27.2 (12.2, 42.2), p = 0.002 | -0.883 (-8.74, 6.97), p = 0.821 |
| Sexuality | 31.1 (7.59, 54.6), p = 0.033 | 26.7 (4.97, 48.4), p = 0.035 | -4.44 (-24.2, 15.3), p = 0.651 |
| Problems with shoulder | -3.96 (-14.9, 7), p = 1.000 | 0.833 (-3.76, 5.43), p = 1.000 | 4.79 (-7.87, 17.5), p = 1.000 |
| Skin Problems | 37 (20.8, 53.2), p = <0.001 | 21.1 (7.87, 34.4), p = 0.005 | -15.9 (-36.1, 4.28), p = 0.118 |
| Fear of progression | -3.41 (-17.2, 10.3), p = 0.789 | 7.5 (-10.2, 25.2), p = 0.789 | 10.9 (-1.26, 23.1), p = 0.232 |
| Problems opening mouth | 41 (26.1, 55.8), p = <0.001 | 20 (8.54, 31.5), p = 0.002 | -21 (-33.9, -8.03), p = 0.002 |
| Coughing | 41.3 (23.3, 59.2), p = <0.001 | 10 (-4.88, 24.9), p = 0.181 | -31.3 (-52.6, -9.93), p = 0.011 |
| Social contact | 46.8 (24.1, 69.4), p = <0.001 | 18.3 (1.63, 35), p = 0.032 | -28.4 (-46.6, -10.3), p = 0.006 |
| Swelling in the neck | -24 (-43.5, -4.5), p = 0.052 | -5 (-24.9, 14.9), p = 0.613 | 19 (-0.376, 38.4), p = 0.109 |
| Weight loss | 27 (15.2, 38.8), p = <0.001 | 21.7 (5.11, 38.2), p = 0.024 | -5.29 (-17.3, 6.69), p = 0.375 |
| Problems with wound healing | 12.4 (-3.7, 28.6), p = 0.253 | 13.3 (-1.79, 28.5), p = 0.246 | 0.896 (-11.7, 13.5), p = 0.886 |

| | | | |
|---|---|---|---|
| Neurological problems | 2.93 (-12.9, 18.8), p = 1.000 | 3.33 (-3.46, 10.1), p = 0.977 | 0.405 (-17.1, 17.9), p = 1.000 |

$\Delta\dot{V}O_2/\Delta WR$: V̇O2-work rate relationship, Head and Neck Module 43 items, FEV1: forced expiratory volume in 1 second, FVC: forced vital capacity, HR: heart rate, MVPA: moderate-to vigorous-intensity physical activity, OUES: oxygen uptake efficiency slope, PEF: peak expiratory flow, QLQ-C30: Quality of Life Questionnaire Core 30, RER: respiratory exchange ratio, RPE: rating of perceived exertion, V̇E: minute ventilation, V̇O2 at AT: oxygen consumption at the anaerobic threshold; V̇O2peak: peak oxygen uptake.
