## Supplementary figures and images for "Changes in Cardiorespiratory Fitness in Patients with Human Papillomavirus (HPV)-Related Oropharyngeal Cancer Undergoing Chemoradiotherapy"

### FigA1

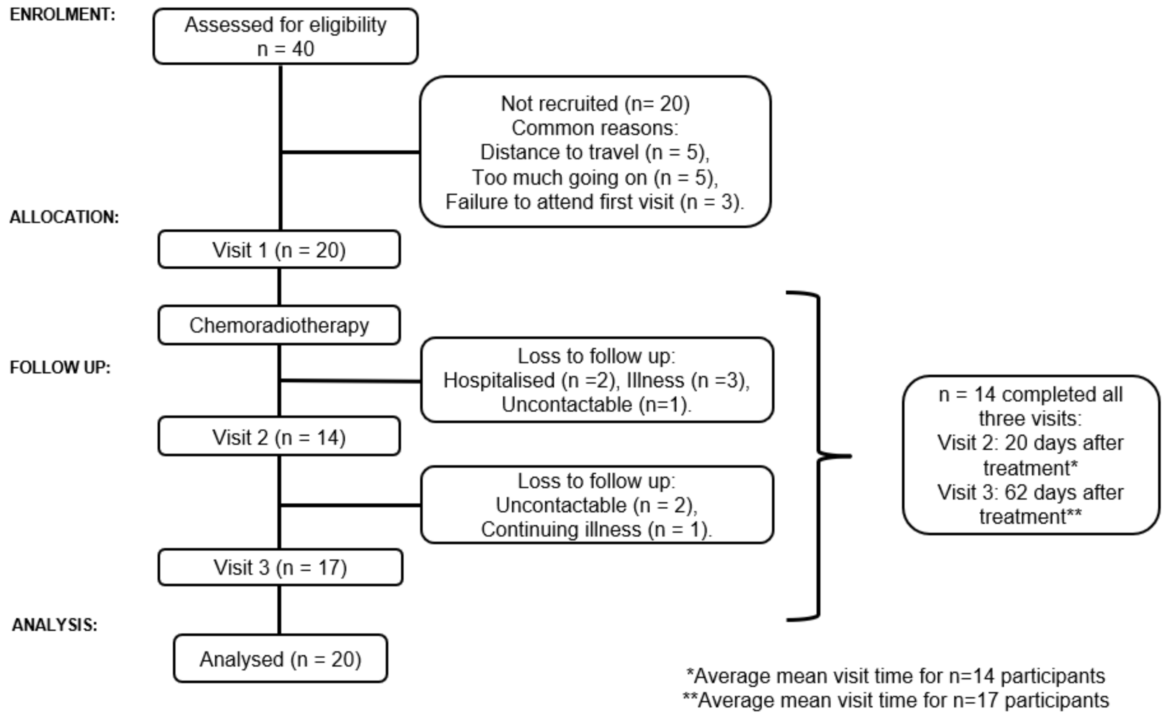


**Figure A1. Participant flow through the study**
